# Prospective and detailed behavioral phenotyping in DDX3X syndrome

**DOI:** 10.1101/2021.01.26.21250125

**Authors:** Lara Tang, Tess Levy, Sylvia Guillory, Danielle Halpern, Jessica Zweifach, Ivy Giserman Kiss, Jennifer H Foss-Feig, Yitzchak Frank, Reymundo Lozano, Puneet Belani, Christina Layton, Emanuel Frowner, Michael S. Breen, Silvia De Rubeis, Ana Kostic, Alexander Kolevzon, Joseph D Buxbaum, Paige M Siper, Dorothy E Grice

## Abstract

**Background:** DDX3X syndrome is a recently identified genetic disorder that accounts for 1-3% of cases of unexplained developmental delay (DD) and/or intellectual disability (ID) in females and is associated with motor and language delays, and autism spectrum disorder (ASD). To date, the published phenotypic characterization of this syndrome has primarily relied on medical record review; in addition, the behavioral dimensions of the syndrome have not been fully explored.

**Methods:** We carried out multi-day, prospective, detailed phenotyping of DDX3X syndrome in 14 females and 1 male, focusing on behavioral, psychological, and neurological measures; three participants in this cohort have been previously reported. We compared results against population norms and contrasted phenotypes between individuals harboring either (i) protein-truncating variants or (ii) missense variants and in-frame deletions.

**Results:** Eighty percent of individuals met criteria for ID, 60% for ASD and 53% for attention-deficit/hyperactivity disorder (ADHD). Motor and language delays were common as were sensory processing abnormalities. The cohort included 5 missense, 3 intronic/splice-site, 2 nonsense, 2 frameshift, 2 in-frame deletions, and one initiation codon variant. Genotype-phenotype correlations indicated that missense variants/in-frame deletions were associated with more severe language, motor, and adaptive deficits in comparison to protein-truncating variants.

**Limitations:** Sample size is modest, however, DDX3X is a rare and underdiagnosed disorder.

**Conclusion:** This study, representing a first, prospective, detailed characterization of DDX3X syndrome, extends our understanding of the neurobehavioral phenotype. Gold-standard diagnostic approaches demonstrated high rates of ID, ASD, and ADHD. In addition, sensory deficits were observed to be a key part of the syndrome. Even with a modest sample, we observe evidence for genotype-phenotype correlations with missense variants/in-frame deletions yielding a more severe phenotype.

## BACKGROUND

A significant proportion of ASD and associated neurodevelopmental disorders (NDDs), including ID, global developmental delay (DD) and epilepsy, are linked to ultrarare genetic variants. As exome and genome sequencing become more accessible, many novel genetic NDD syndromes are being identified. These include DDX3X syndrome, which is emerging as one of the most common genetic causes of ID/DD in females [1, 2] and is also significantly associated with ASD [3-6].

The *DDX3X* gene encodes a ubiquitously expressed ATP-dependent DEAD-box RNA helicase, involved in mRNA biogenesis, RNA metabolism, and mRNA translation [7, 8]. Recently, it has been shown that DDX3X is involved in the anti-viral innate immune response, stress granule nucleation and localization, apoptotic signaling following DNA damage, maintenance of lipid homeostasis, cell cycle control, and regulation of Wnt-β-catenin signaling [8-11]. In the central nervous system, DDX3X is essential for neurite outgrowth and synaptogenesis, and for the proliferation and differentiation of cortical neural progenitors [12, 13].

In the large national Deciphering Developmental Disabilities study, which serially ascertained participants for DD, investigators found high rates of *DDX3X* variants (P<10^−50^) [2]. Similarly, a recent large-scale sequencing study of participants ascertained for ASD identified *DDX3X* as a genome-wide significant ASD gene [3], and multiple exome studies also identified *DDX3X* variants in ASD cohorts [3, 5, 6, 14]. Finally, studies of cohorts with *DDX3X* variants demonstrated high rates of developmental delays or intellectual disability (∼50-100%), structural brain changes (25-90%), most commonly, abnormalities of the corpus callosum (25-87%) and cortical dysplasias (11-12%), and behavioral abnormalities, including ASD, hyperactivity, and aggression (21-53% combined) [13, 15-17]. However, these phenotypic studies relied almost exclusively on retrospective data analyses.

*DDX3X* is located in a short region of Xp11.4 that escapes X-inactivation [18, 19], thus females are functionally heterozygous for variants. Most of the identified *DDX3X* variants are in females; it is assumed that males carrying a very deleterious *DDX3X* variant do not survive to term, a finding confirmed in mouse models [18]. Surviving male patients carry missense variants that are thought to act as hypomorphic alleles [20, 21]. Because *DDX3X* variants appear to be very penetrant, all variants in females that have been identified to date are *de novo*.

In the current study, we carried out comprehensive, in-person, prospective phenotyping of 15 participants with DDX3X syndrome, focusing on neurobehavioral outcomes. We compared all normed assessments to population norms, and compared phenotypes in participants with protein-truncating variants versus other variants.

## METHODS

Fifteen individuals (14 female;1 male) from 3 to 16 years old (7.5±4.5 years) diagnosed with DDX3X syndrome were evaluated at the Seaver Autism Center at the Icahn School of Medicine at Mount Sinai (Figure 1). Three participants have been previously reported (Table S1). All participants were seen for a 3 to 4 day in-person visit at the Seaver Center, including approximately 9 hours of direct assessment, 6 hours of caregiver interviews, and 4 hours of caregiver questionnaires. All neuropsychological testing was completed by research-reliable clinical psychologists. Medical evaluations (psychiatric, neurologic, and clinical genetic) were completed by board-certified clinicians (Table 1). In addition, a board-certified radiologist, with specialty training in neuroradiology, reviewed all available MRI scans. All other available medical records were reviewed by the study lead. This study was approved by the Mount Sinai Institutional Review Board. Legal guardians of each participant gave informed consent prior to study participation and additional consent prior to publication.

**Table 1:**
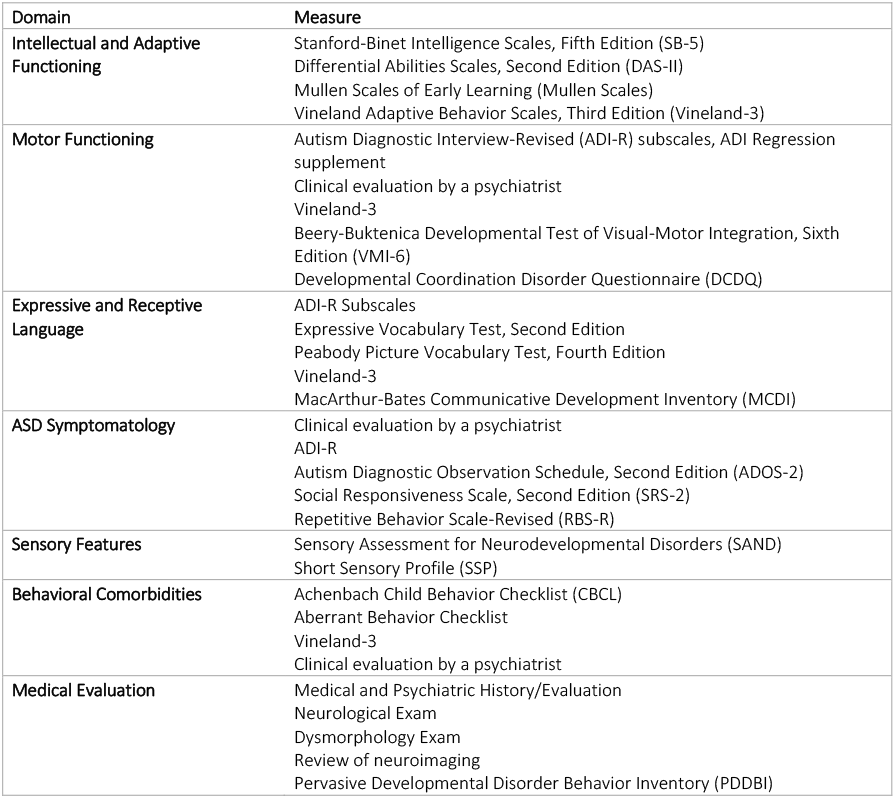
Study Approach.

| Domain | Measure |
| --- | --- |
| **Intellectual and Adaptive Functioning** | Stanford-Binet Intelligence Scales, Fifth Edition (SB-5)<br>Differential Abilities Scales, Second Edition (DAS-II)<br>Mullen Scales of Early Learning (Mullen Scales)<br>Vineland Adaptive Behavior Scales, Third Edition (Vineland-3) |
| **Motor Functioning** | Autism Diagnostic Interview-Revised (ADI-R) subscales, ADI Regression supplement<br>Clinical evaluation by a psychiatrist<br>Vineland-3<br>Beery-Buktenica Developmental Test of Visual-Motor Integration, Sixth Edition (VMI-6)<br>Developmental Coordination Disorder Questionnaire (DCDQ) |
| **Expressive and Receptive Language** | ADI-R Subscales<br>Expressive Vocabulary Test, Second Edition<br>Peabody Picture Vocabulary Test, Fourth Edition<br>Vineland-3<br>MacArthur-Bates Communicative Development Inventory (MCDI) |
| **ASD Symptomatology** | Clinical evaluation by a psychiatrist<br>ADI-R<br>Autism Diagnostic Observation Schedule, Second Edition (ADOS-2)<br>Social Responsiveness Scale, Second Edition (SRS-2)<br>Repetitive Behavior Scale-Revised (RBS-R) |
| **Sensory Features** | Sensory Assessment for Neurodevelopmental Disorders (SAND)<br>Short Sensory Profile (SSP) |
| **Behavioral Comorbidities** | Achenbach Child Behavior Checklist (CBCL)<br>Aberrant Behavior Checklist<br>Vineland-3<br>Clinical evaluation by a psychiatrist |
| **Medical Evaluation** | Medical and Psychiatric History/Evaluation<br>Neurological Exam<br>Dysmorphology Exam<br>Review of neuroimaging<br>Pervasive Developmental Disorder Behavior Inventory (PDDBI) |

**Figure 1.**
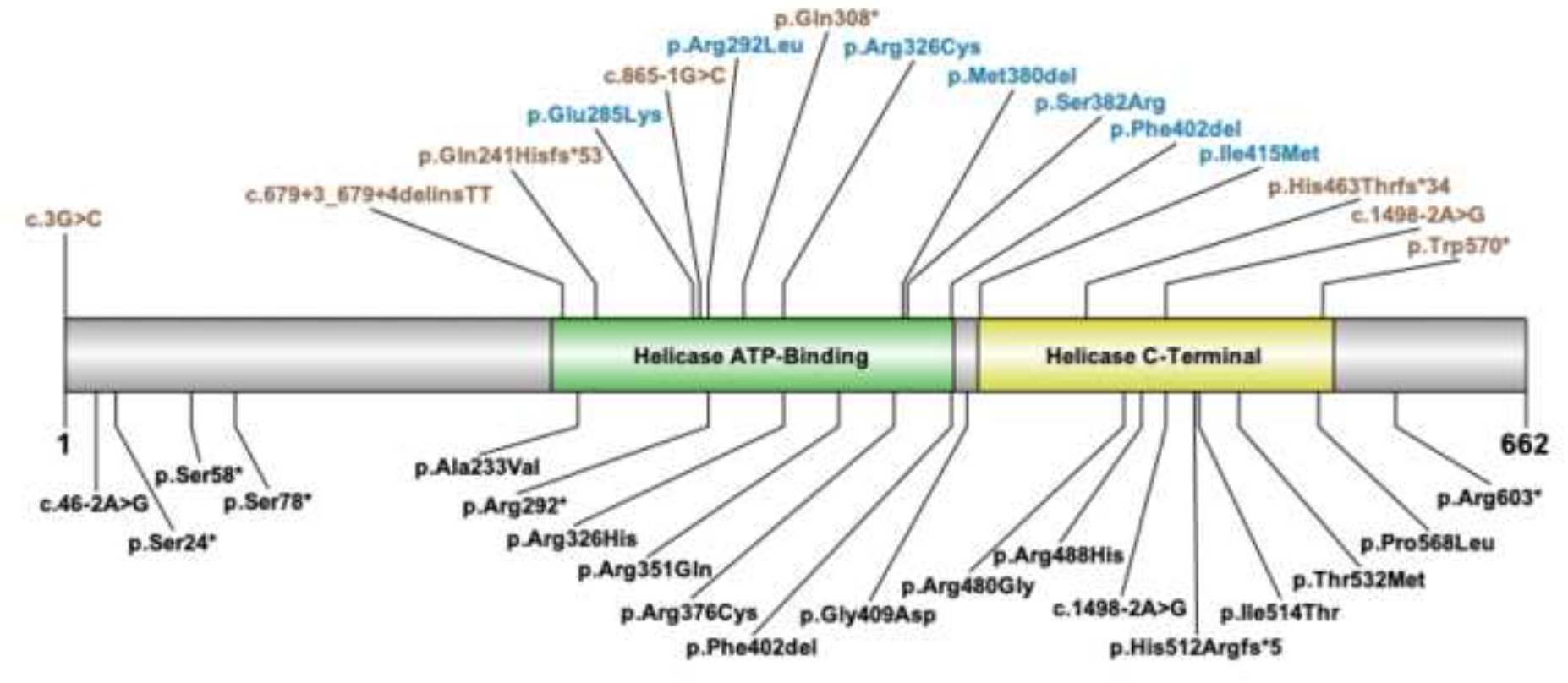
DDX3X variants. Top, variants in the cohort: Protein-truncating variants (PTVs) are colored tan, while missense variants and in-frame deletions are colored blue. The male participant carries the p.Arg292Leu variant. Bottom, recurrent variants: Variants reported at least three times in the literature and/or Clinvar. The helicase ATP-binding and helicase C-terminal domains are shown as reported in Uniprot O00571.

### Genetic Testing

Variants were identified and validated at Clinical Laboratory Improvement Amendments certified laboratories, as was *de novo* status. Variants were reannotated according to the Human Genome Variation Society Guidelines, with nucleotide and amino acid positions mapped to the DDX3X RefSeq transcript NM_001356.4, and interpreted using the American College of Medical Genetics and Genomics and Association for Molecular Pathology Guidelines [22] (Table S1). Variants were annotated as either (i) protein-truncating variants (PTVs, including nonsense, frameshift, splice-site and start-codon loss variants), or (ii) missense or in-frame deletions.

### Intellectual and Adaptive Function

Intellectual functioning was assessed by clinical psychologists using the Stanford-Binet Intelligence Scales, 5^th^ Edition (SB-5) [23], the Differential Abilities Scales, 2^nd^ Edition (DAS-II) [24], or the Mullen Scales of Early Learning (Mullen Scales) [25]. Full Scale Intelligence Quotient (IQ), nonverbal IQ and verbal IQ scores were calculated for those participants who completed the SB-5 or the DAS-II. Developmental quotient (DQ) scores were calculated from age equivalents for all participants and used for a cohort-wide comparison of cognitive abilities. Adaptive functioning was evaluated using the Vineland Adaptive Behavior Scales, 3^rd^ Edition (Vineland-3) [26], which was administered by clinical psychologists. ID was diagnosed based on the Diagnostic and Statistical Manual of Mental Disorders, Fifth Edition (DSM-5) criteria [27].

### Motor Function

Major motor milestones were evaluated using the Autism Diagnostic Interview-Revised (ADI-R) [28] and ADI regression supplement [29], administered by clinical psychologists, and psychiatric evaluation, by a child and adolescent psychiatrist. Fine and gross motor skills were evaluated using the Vineland-3 and Mullen subscales. To assess the integration of skills in the visual-motor domain, clinical psychologists administered the Beery-Buktenica Developmental Test of Visual-Motor Integration, Sixth Edition (VMI-6) [30]. Caregivers completed the Developmental Coordination Disorder Questionnaire (DCDQ) [31].

### Expressive and Receptive Language

Language milestones were obtained using the ADI-R and the ADI regression supplement. Direct assessment of expressive language was assessed by the Expressive Vocabulary Test, Second Edition [32], and receptive language by the Peabody Picture Vocabulary Test, Fourth Edition [33], both administered by clinical psychologists. The language subdomains of the Vineland-3 and Mullen Scales were also analyzed. Caregivers completed the MacArthur-Bates Communicative Development Inventory (MCDI) [34].

### ASD Symptomatology and Sensory Features

Consensus DSM-5 [27] ASD diagnoses were determined from the psychiatric evaluation and results from gold-standard diagnostic assessments including the Autism Diagnostic Observation Schedule, Second Edition (ADOS-2) [35], and the ADI-R [28]. Caregiver questionnaires, including the Social Responsiveness Scale, Second Edition (SRS-2) [36] and the Repetitive Behavior Scale-Revised (RBS-R) [37], were used to further evaluate ASD features.

Sensory symptoms were assessed using the Sensory Assessment for Neurodevelopmental Disorders (SAND) [38], administered by a clinical psychologist, and the Short Sensory Profile (SSP) [39], a caregiver questionnaire.

### Behavioral Comorbidities

The Achenbach Child Behavior Checklist (CBCL) [40] and the Aberrant Behavior Checklist [41-44] were used to assess additional psychiatric features and behavioral challenges. The Vineland-3 Maladaptive Behavior domain and subdomains of Internalizing and Externalizing Behavior were evaluated. Other behavioral and psychiatric comorbidities were documented during the psychiatric evaluation.

### Medical Evaluation

A neurological examination was completed by a pediatric neurologist. The examination assessed motor and sensory skills, balance and coordination, mental status, and reflexes. Dysmorphic features were assessed by a clinical geneticist. Medical history was assessed by parent report and review of medical records by the study psychiatrist. Brain MRI scans (n=12) or clinical reports (n=2) were reviewed by the study neuroradiologist, and clinically significant findings documented. Caregivers completed the Pervasive Developmental Disorder Behavior Inventory (PDDBI) [45] to supplement characterization of sleep disturbance and prenatal and neonatal complications.

## RESULTS

All *DDX3X* variants were classified as pathogenic or likely pathogenic. Variants included 5 missense variants, 3 intronic/splice-site variants, 2 nonsense variants, 2 frameshift variants, 2 in-frame deletions, and one initiation codon variant (Figure 1, Table S1). The variants clustered in the helicase ATP-Binding and helicase C-Terminal domains. *De novo* status was confirmed in 14/15 individuals, and the remaining individual had maternal inheritance ruled out. The male participant carried a *de novo* missense variant.

### Intellectual and Adaptive Functioning

Nine of the 15 participants in the cohort completed the Mullen Scales, 5 completed the SB-5, and 1 was administered the DAS-II. Standard scores (population M=100, SD=15) across the 6 individuals for which IQs could be calculated ranged from 40 to 85 (58.7±17.2) for full scale IQ, 42 to 87 (59.7±16.1) for nonverbal IQ, and 43 to 111 (65.7±25.6) for verbal IQ. Full scale DQ, nonverbal DQ, and verbal DQ were calculated for all participants: 14/15 participants showed greater than a 40% delay in all scores; one participant showed average scores for full scale DQ, nonverbal DQ, and an above average score in verbal DQ (Table 2, Figure 2A). On the Vineland-3, average standard scores for Communication, Daily Living Skills, Socialization, and the Adaptive Behavior Composite were 3 to 4 standard deviations below the general population mean (Table 2, Figure 2B, Table S2).

**Table 2:**
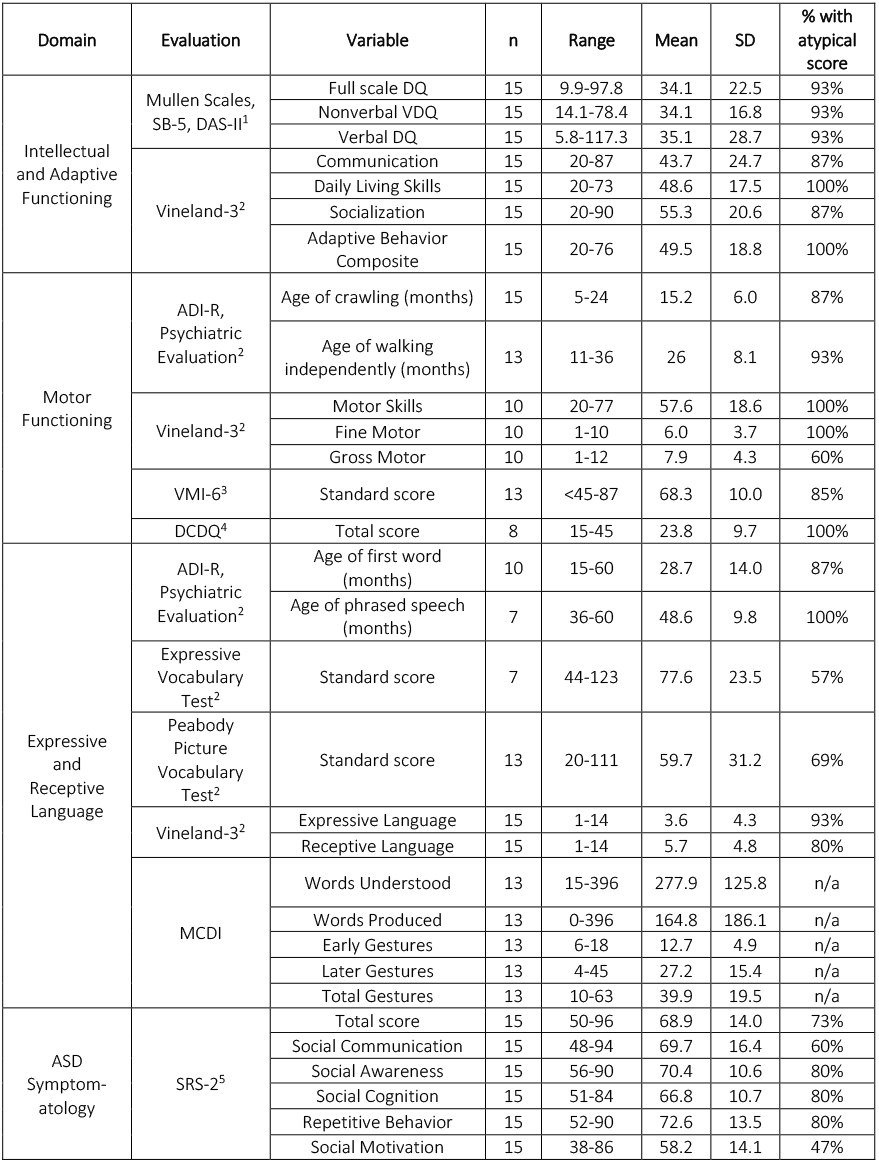
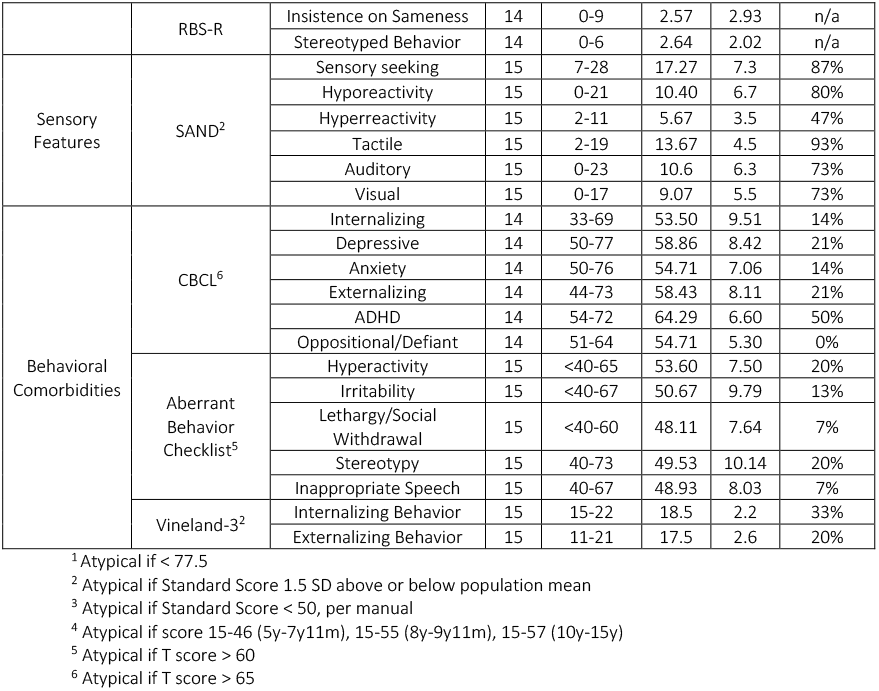
Summary statistics for clinical measures.

| Domain | Evaluation | Variable | n | Range | Mean | SD | % with atypical score |
| --- | --- | --- | --- | --- | --- | --- | --- |
| Intellectual and Adaptive Functioning | Mullen Scales, SB-5, DAS-II<sup>1</sup> | Full scale DQ | 15 | 9.9-97.8 | 34.1 | 22.5 | 93% |
|  |  | Nonverbal VDQ | 15 | 14.1-78.4 | 34.1 | 16.8 | 93% |
|  |  | Verbal DQ | 15 | 5.8-117.3 | 35.1 | 28.7 | 93% |
|  | Vineland-3<sup>2</sup> | Communication | 15 | 20-87 | 43.7 | 24.7 | 87% |
|  |  | Daily Living Skills | 15 | 20-73 | 48.6 | 17.5 | 100% |
|  |  | Socialization | 15 | 20-90 | 55.3 | 20.6 | 87% |
|  |  | Adaptive Behavior Composite | 15 | 20-76 | 49.5 | 18.8 | 100% |
| Motor Functioning | ADI-R, Psychiatric Evaluation<sup>2</sup> | Age of crawling (months) | 15 | 5-24 | 15.2 | 6.0 | 87% |
|  |  | Age of walking independently (months) | 13 | 11-36 | 26 | 8.1 | 93% |
|  | Vineland-3<sup>2</sup> | Motor Skills | 10 | 20-77 | 57.6 | 18.6 | 100% |
|  |  | Fine Motor | 10 | 1-10 | 6.0 | 3.7 | 100% |
|  |  | Gross Motor | 10 | 1-12 | 7.9 | 4.3 | 60% |
|  | VMI-6<sup>3</sup> | Standard score | 13 | <45-87 | 68.3 | 10.0 | 85% |
|  | DCDQ<sup>4</sup> | Total score | 8 | 15-45 | 23.8 | 9.7 | 100% |
| Expressive and Receptive Language | ADI-R, Psychiatric Evaluation<sup>2</sup> | Age of first word (months) | 10 | 15-60 | 28.7 | 14.0 | 87% |
|  |  | Age of phrased speech (months) | 7 | 36-60 | 48.6 | 9.8 | 100% |
|  | Expressive Vocabulary Test<sup>2</sup> | Standard score | 7 | 44-123 | 77.6 | 23.5 | 57% |
|  | Peabody Picture Vocabulary Test<sup>2</sup> | Standard score | 13 | 20-111 | 59.7 | 31.2 | 69% |
|  | Vineland-3<sup>2</sup> | Expressive Language | 15 | 1-14 | 3.6 | 4.3 | 93% |
|  |  | Receptive Language | 15 | 1-14 | 5.7 | 4.8 | 80% |
|  | MCDI | Words Understood | 13 | 15-396 | 277.9 | 125.8 | n/a |
|  |  | Words Produced | 13 | 0-396 | 164.8 | 186.1 | n/a |
|  |  | Early Gestures | 13 | 6-18 | 12.7 | 4.9 | n/a |
|  |  | Later Gestures | 13 | 4-45 | 27.2 | 15.4 | n/a |
|  |  | Total Gestures | 13 | 10-63 | 39.9 | 19.5 | n/a |
| ASD Symptomatology | SRS-2<sup>5</sup> | Total score | 15 | 50-96 | 68.9 | 14.0 | 73% |
|  |  | Social Communication | 15 | 48-94 | 69.7 | 16.4 | 60% |
|  |  | Social Awareness | 15 | 56-90 | 70.4 | 10.6 | 80% |
|  |  | Social Cognition | 15 | 51-84 | 66.8 | 10.7 | 80% |
|  |  | Repetitive Behavior | 15 | 52-90 | 72.6 | 13.5 | 80% |
|  |  | Social Motivation | 15 | 38-86 | 58.2 | 14.1 | 47% |
|  | RBS-R | Insistence on Sameness | 14 | 0-9 | 2.57 | 2.93 | n/a |
|  |  | Stereotyped Behavior | 14 | 0-6 | 2.64 | 2.02 | n/a |
| Sensory Features | SAND<sup>2</sup> | Sensory seeking | 15 | 7-28 | 17.27 | 7.3 | 87% |
|  |  | Hyporeactivity | 15 | 0-21 | 10.40 | 6.7 | 80% |
|  |  | Hyperreactivity | 15 | 2-11 | 5.67 | 3.5 | 47% |
|  |  | Tactile | 15 | 2-19 | 13.67 | 4.5 | 93% |
|  |  | Auditory | 15 | 0-23 | 10.6 | 6.3 | 73% |
|  |  | Visual | 15 | 0-17 | 9.07 | 5.5 | 73% |
| Behavioral Comorbidities | CBCL<sup>6</sup> | Internalizing | 14 | 33-69 | 53.50 | 9.51 | 14% |
|  |  | Depressive | 14 | 50-77 | 58.86 | 8.42 | 21% |
|  |  | Anxiety | 14 | 50-76 | 54.71 | 7.06 | 14% |
|  |  | Externalizing | 14 | 44-73 | 58.43 | 8.11 | 21% |
|  |  | ADHD | 14 | 54-72 | 64.29 | 6.60 | 50% |
|  |  | Oppositional/Defiant | 14 | 51-64 | 54.71 | 5.30 | 0% |
|  | Aberrant Behavior Checklist<sup>5</sup> | Hyperactivity | 15 | <40-65 | 53.60 | 7.50 | 20% |
|  |  | Irritability | 15 | <40-67 | 50.67 | 9.79 | 13% |
|  |  | Lethargy/Social Withdrawal | 15 | <40-60 | 48.11 | 7.64 | 7% |
|  |  | Stereotypy | 15 | 40-73 | 49.53 | 10.14 | 20% |
|  |  | Inappropriate Speech | 15 | 40-67 | 48.93 | 8.03 | 7% |
|  | Vineland-3<sup>2</sup> | Internalizing Behavior | 15 | 15-22 | 18.5 | 2.2 | 33% |
|  |  | Externalizing Behavior | 15 | 11-21 | 17.5 | 2.6 | 20% |

<sup>1</sup> Atypical if < 77.5
<sup>2</sup> Atypical if Standard Score 1.5 SD above or below population mean
<sup>3</sup> Atypical if Standard Score < 50, per manual
<sup>4</sup> Atypical if score 15-46 (5y-7y11m), 15-55 (8y-9y11m), 15-57 (10y-15y)
<sup>5</sup> Atypical if T score > 60
<sup>6</sup> Atypical if T score > 65

**Figure 2.**
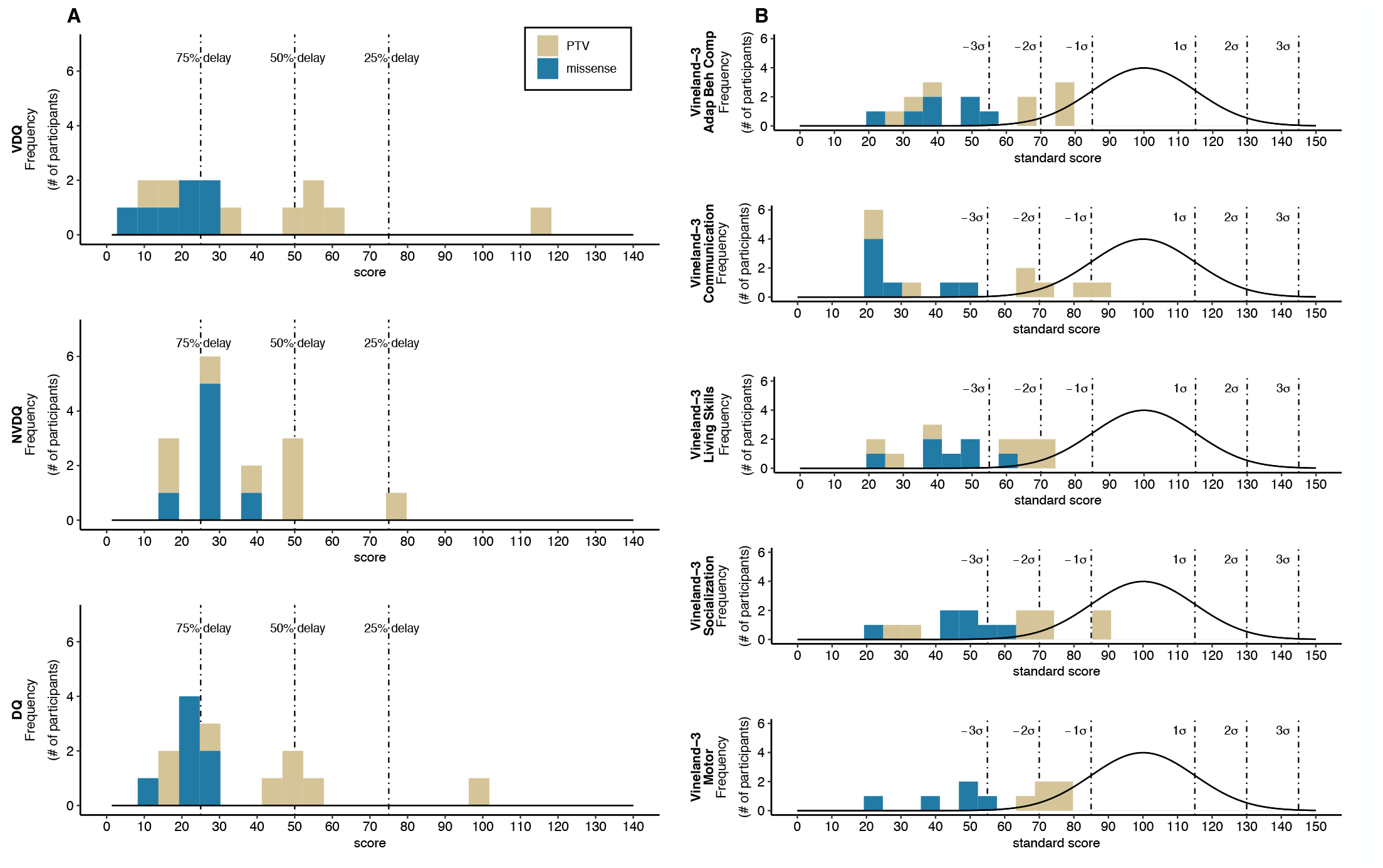
Intellectual, adaptive and motor functioning. A. Frequency histograms of verbal, nonverbal, and full-scale developmental quotients (VDQ, NVDQ, DQ). B. Frequency histograms of standard scores on domains of the Vineland-3: Adaptive Behavior Composite, Communication, Daily Living Skills, Socialization, and Motor (the Maladaptive Behavior domain is represented in Supplemental Figure 1B). All plots show frequency (i.e., number of individuals) in each bin. Developmental delays of 25%, 50%, and 75% are indicated by dashed lines (A), while distribution of standard scores in typically developing individuals are shown as black lines (B), together with associated standard deviations (dashed lines). PTV, protein truncating variant; missense, missense variants or in-frame deletions.

Overall, 12 of 15 participants in this cohort were given a diagnosis of ID based on DSM-5 criteria.

### Motor Functioning

Delays in age of first crawling were reported in 13 of 15 participants (Table 2). Two of the participants, both 5 years old, could not walk unaided at the time of assessment, while age of first walking in the 13 other participants ranged from 11 to 36 months.

Ten participants in the cohort were administered the Motor domain of the Vineland-3 (Figure 2B, Table S2). Average scores in the Motor Domain and Gross Motor subdomain were 3 standard deviations below the general population mean, while the Fine Motor subdomain scores were 4 standard deviations below the mean (Table 2). Based on the Vineland-3, 2 individuals had significantly higher gross motor than fine motor ability, while 1 showed higher fine motor ability. On the Mullen Scales, 4 individuals showed higher gross motor than fine motor abilities. However, overall the cohort did not show significant differences between fine and gross motor ability. On the VMI-6 (n=13, 2 attempted but could not complete), the average standard score of the cohort was 2 standard deviations below the general population mean, with 7 individuals scoring in the impaired range (Table S2). On the DCDQ (n=8), 100% of participants scored in a range indicative and/or suspect of a Developmental Coordination Disorder.

### Expressive and Receptive Language

Early language milestones, collected from the ADI-R and psychiatric evaluation, were delayed for all 15 participants. At the time of assessment, 5 participants in the cohort were non-verbal (Figure 3A). Of the 10 participants with verbal capacity, the age when first words emerged ranged from 15 to 60 months, and 7 individuals achieved phrase speech that emerged from 36 to 60 months (Table 2).

**Figure 3.**
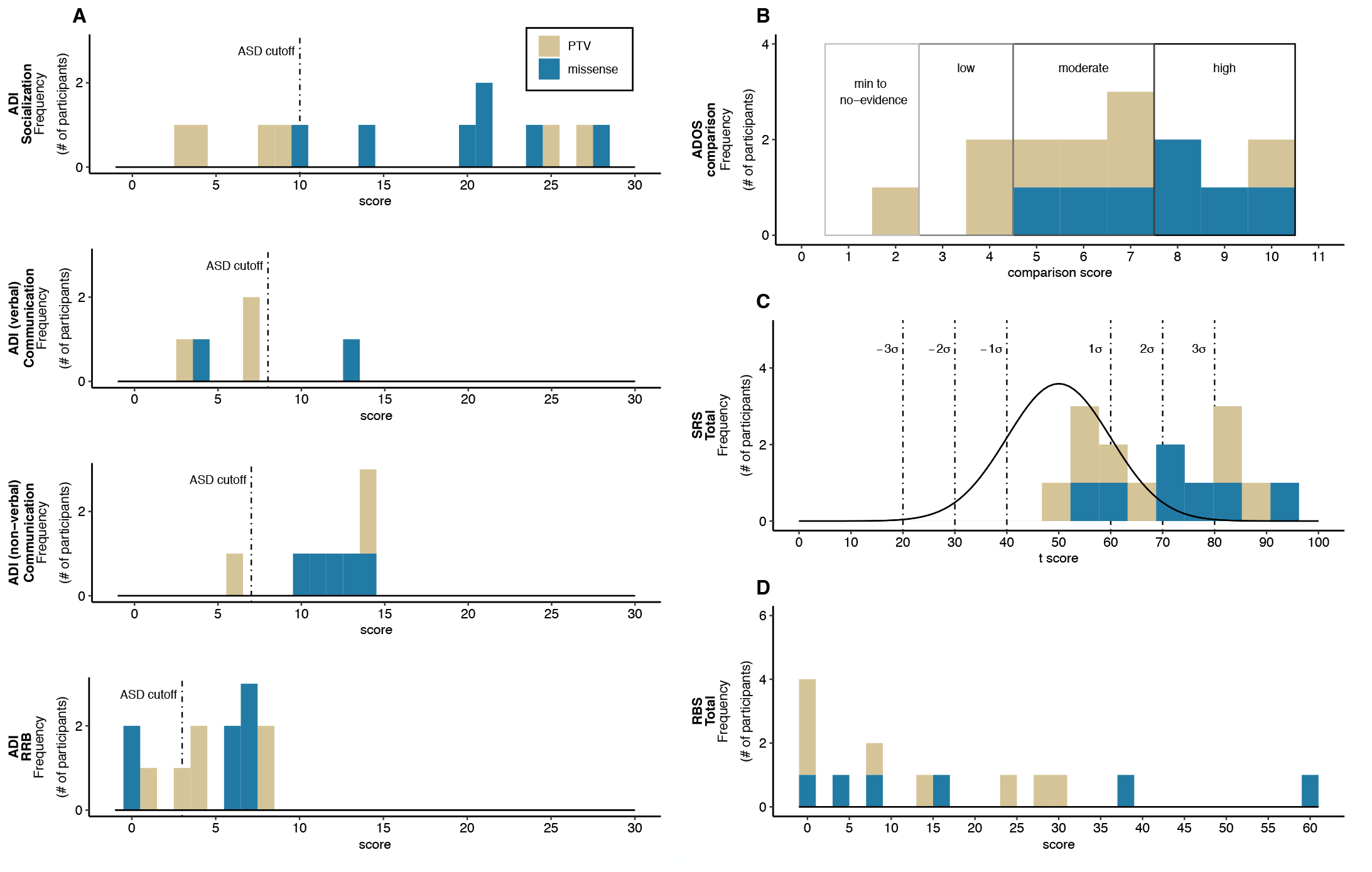
Psychiatric features. A. Frequency histograms for the Autism Diagnostic Interview - Revised (ADI-R) Socialization, Communication, and Restricted/Repetitive Behavior subdomains (RRB). Dashed lines, represent the diagnostic threshold for ASD for each subdomain, with scores to right surpassing the threshold. B. Frequency histograms for the Autism Diagnostic Observation Schedule - Second Edition (ADOS-2) comparison score. Evidence levels for ASD are categorized as minimum to no evidence, low, moderate, and high evidence. C. Frequency histograms for the Social Responsiveness Scale (SRS-2) total score. Total score has a mean of 50 and standard deviation of 10. D. Frequency histograms for the Repetitive Behaviors Scale - Revised (RBS-R). Total scores have a minimum of 0 and maximum of 129. In all plots, higher scores indicate greater deficits. In panels C & D, distribution of standard scores in typically developing individuals are shown as black lines, together with associated standard deviations (dashed lines). PTV, protein truncating variant; missense, missense variants or in-frame deletions.

Expressive language, assessed by the Expressive Vocabulary Test, Second Edition, was completed by 7 participants; 8 did not have skills to achieve the base score. For those who completed the assessment, standard scores ranged from 44 to 123 (77.6±23.5). Receptive language, assessed by the Peabody Picture Vocabulary Test, Fourth Edition, was completed by 13 participants, while 2 did not have the skills to achieve the base score; standard scores ranged from 20 to 111 (59.7±31.2). On the Vineland-3, Expressive and Receptive Language subdomains showed similar variability (Table 2). Receptive and expressive language was compared within individuals using the Expressive Vocabulary Test and Peabody Picture Vocabulary Test (n=7) and the Vineland-3 subscales (n=15). Using the Peabody Picture Vocabulary Test and Expressive Vocabulary Test, 2 individuals showed significantly higher expressive than receptive language ability, and 2 showed significantly higher receptive than expressive ability (>12 difference in standard score). However, it is important to note that 4 individuals could complete the Peabody Picture Vocabulary Test but could not complete the Expressive Vocabulary Test. Using the Vineland-3, 47% showed significantly higher receptive than expressive language ability.

On the MCDI (n=13), parents reported the average number of words understood was 278 and words produced was 165 (from of a total of 396 queried), again indicating higher receptive than expressive skills. On average, individuals had 13 early gestures (e.g., shakes head no, blows kiss), and 27 later gestures (e.g., brushes teeth, waters plants), with the average of total gestures being 40 (from a total of 63).

### ASD Symptomatology

Nine of 15 participants (60%) received a consensus DSM-5 diagnosis of ASD based on clinical evaluation, ADOS-2, and ADI-R.

Individuals in this cohort were administered ADOS-2 Module 1 (n=10), 2 (n=3), 3 (n=1), and 4 (n=1). Based on results from the ADOS-2 alone, 26.7% of participants did not meet diagnostic criteria for autism or autism spectrum classifications, whereas 20% of participants received scores that met the autism spectrum classification and 53.3% of participants received scores that met for an autism classification (Figure 3B, Table S2). The ADOS-2 had 9 true positive results and 3 true negative results, but 3 false positive results, when compared to consensus diagnosis. False positives were present in all 3 participants who received an ADOS-2 Module 2, which is administered to individuals with phrase speech. A review of results from the ADOS-2, and cognitive and language assessments, indicated that ADOS-2 scores in these participants were impacted by the presence of language delays and repetitive behavior domain symptoms (e.g., sensory interests).

Seven of 13 participants met the diagnostic threshold for autism in all four domains on the ADI-R (two caregivers were not administered the ADI-R to reduce caregiver strain once a diagnosis of ASD was excluded) (Figure 3A, Table S2). Examining individual domains, 9 of these 13 participants met or surpassed the cutoff on the Socialization domain, 8 met the cutoff on the Communication domain, 10 met the cutoff on the Restricted and Repetitive Behavior domain, and all individuals who completed the ADI-R met the cutoff on the Abnormality Evident by 36 Months domain.

Results from the SRS-2 (n=15) indicated that 80% of the cohort had deficits in the Social Awareness, Social Cognition, and Repetitive Behavior domains, 60% showed deficits in the Social Communication domain, and 47% in the Social Motivation domain (Figure 3C). On the RBS-R (n=14), the greatest number of symptoms was reported for Insistence on Sameness (36 symptoms reported across the cohort, with individual participant symptom counts ranging from 0 to 9) and Stereotyped Behavior (37 symptoms reported across the cohort, with individual symptom counts ranging from 0 to 6) (Figure 3D).

### Sensory Features

Sensory responsivity data were collected through the SAND (n=15), and z-scores were calculated based on an age-matched sample of typically developing (TD) controls (n=29) (Figure 4). The DDX3X group displayed significantly more sensory symptoms across symptom domains and sensory modalities compared to the TD group (Table 2). Sensory hyperreactivity scores ranged from average to >3 SDs above the mean. The majority of participants scored >2 SDs above the mean in the sensory hyporeactivity domain (12/15) and >2 SDs above the mean in the sensory seeking domain (13/15). On average, sensory seeking and hyporeactivity were more common than hyperreactivity symptoms. Within sensory modalities, the cohort showed a greater number of tactile symptoms compared to auditory and visual symptoms, although scores were elevated relative to controls in all modalities. Tactile hyporeactivity (e.g., high pain threshold) was more commonly observed than visual and auditory hyporeactivity, while visual hyperreactivity was more common than tactile and auditory hyperreactivity.

**Figure 4.**
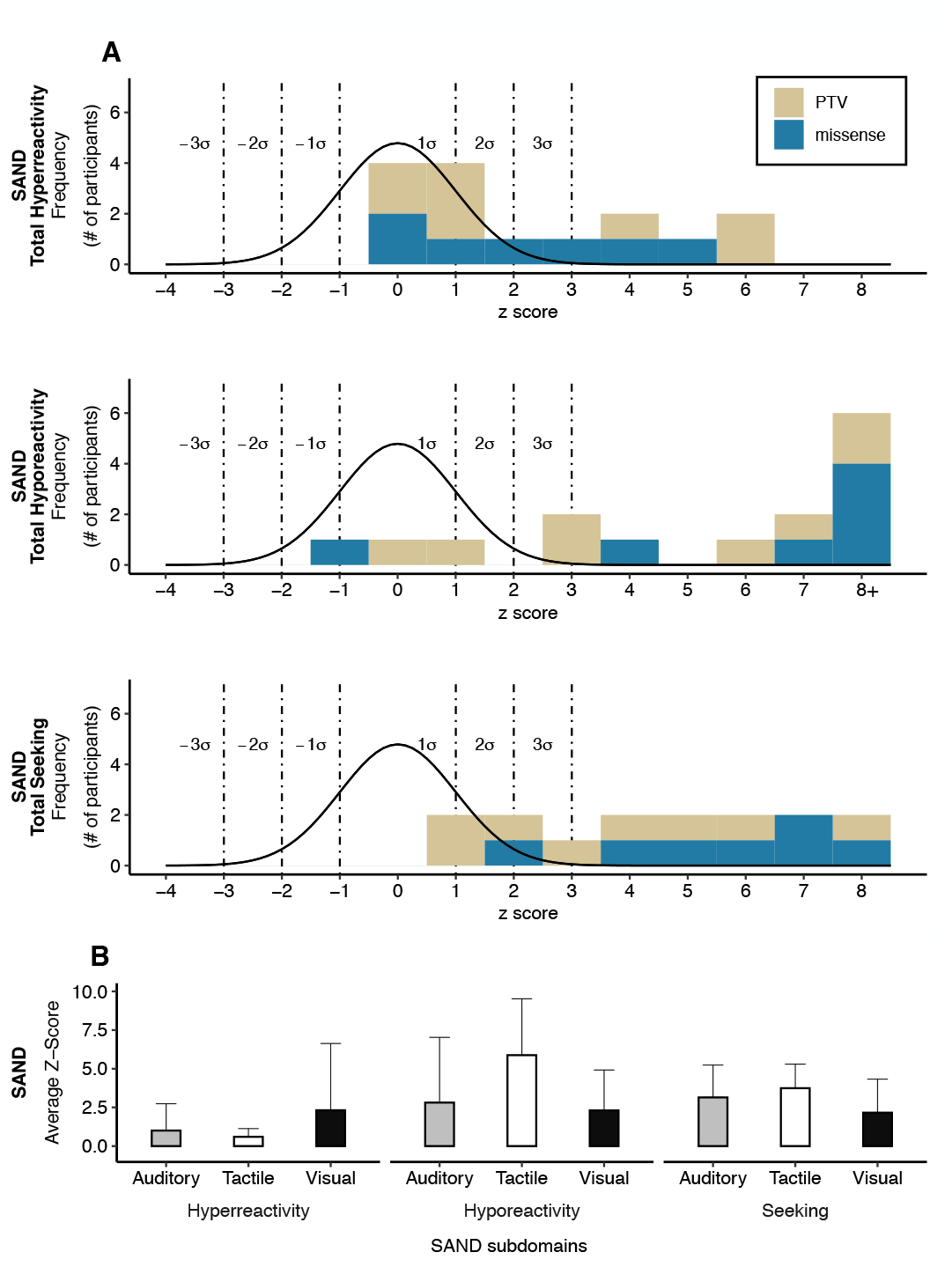
Sensory reactivity. A. Frequency histograms for the Sensory Assessment for Neurodevelopmental Disorders (SAND) hyperreactivity, hyporeactivity and seeking domains. Distribution of standard scores in typically developing individuals are shown as black lines, together with associated standard deviations (dashed lines). B. Average Z-scores for hyperreactivity, hyporeactivity and seeking within visual, tactile, and auditory modalities. Z-scores have a mean of 0 where +1 indicates 1 SD above the mean. PTV, protein truncating variant; missense, missense variants or in-frame deletions.

On the SSP (n=15), total scores indicated definite sensory differences in 40% of individuals, possible differences in 26.7% individuals, and typical performance in 33.3% individuals. The greatest number of reported symptoms was in the Under-responsive/Seeks Sensation subdomain, in which definite sensory differences were reported in 11 individuals.

### Behavioral Comorbidities

Domains from the school-age and pre-school versions of the CBCL (n=14) were combined to evaluate additional psychiatric symptomatology (Table 2, Figure S1-A). Results from the Internalizing domain indicated clinically significant results for 14% of the cohort and results from the Externalizing domain indicated clinically significant results in 21% of the cohort.

Results from the Aberrant Behavior Checklist indicated that 20% of the cohort scored in the clinically significant range in the Hyperactivity and Stereotypy domains, 13% in the Irritability domain, and 7% in each the Lethargy/Social Withdrawal and Inappropriate Speech domains.

On the Internalizing subdomain of the Maladaptive Behavior domain from the Vineland-3, v-scores ranged from 15 to 22 (18.5±2.2) and from 11 to 21 (17.5±2.6) on the Externalizing subdomain (Figure S1-B). Clinical evaluation by the psychiatrist and review of all available assessments indicated that 8 of 15 participants met consensus diagnoses for ADHD, combined type, and 1 for Generalized Anxiety Disorder.

### Medical Evaluation

Medical, neurological and clinical genetic findings are separated by frequency, operationalized as frequent (>50%), common (20-50%), and other less frequent findings (<20%) (Table 3 and Tables S3-S5).

**Table 3:** Common (>50%) medical findings, including dysmorphisms

| Feature | Percentage | n |
| --- | --- | --- |
| Gait abnormalities | 100% | 15/15 |
| Hypotonia | 93% | 14/15 |
| Malar hypoplasia | 93% | 13/14 |
| Flat midface | 93% | 13/14 |
| Structural brain changes | 86% | 12/14 |
| Corpus callosum abnormalities | 64% | 9/14 |
| Key hole shaped temporal horns | 50% | 7/14 |
| Enlarged ventricles | 43% | 6/14 |
| White matter abnormalities | 43% | 6/14 |
| Vermis abnormalities | 21% | 3/14 |
| Gastrointestinal problems | 80% | 12/15 |
| Constipation | 53% | 8/15 |
| Reflux | 53% | 8/15 |
| Ocular abnormalities | 73% | 11/15 |
| Strabismus | 40% | 6/15 |
| Astigmatism | 33% | 5/15 |
| Myopia | 20% | 3/15 |
| Amblyopia | 13% | 2/15 |
| Hyperopia | 13% | 2/15 |
| Sleep disturbance | 67% | 10/15 |
| Feeding issues | 67% | 10/15 |
| Hyperextensibility | 64% | 9/14 |
| Pigmented skin lesions | 62% | 8/13 |
| Bulbous nose | 57% | 8/14 |
| High arched palate | 57% | 8/14 |
| Recurrent infections | 53% | 8/15 |
| Otitis media | 40% | 6/15 |
| Urinary tract | 20% | 3/15 |
| Respiratory tract | 20% | 3/15 |
| Pointed chin | 50% | 7/14 |

#### Frequent findings

Gait disturbance was ubiquitous (15/15). Gait abnormalities include apraxic, ataxic, or disorganized gait and toe-walking (Table 3, S3-4). Two individuals required walkers. Hypotonia was present in 14/15 and most often considered mild at the time of assessment. Structural brain changes were present in 12 of 14 participants, with the most common finding being an abnormality of the corpus callosum (9/14), followed by key-hole shaped temporal horns (7/14), enlarged ventricles (6/14), white matter abnormalities (6/14), and vermis abnormalities (3/14). One individual had polymicrogyria (PMG). Gastrointestinal problems were frequently reported (12/15); constipation was the most common issue (8/15) followed by gastroesophageal reflux (7/15). Ocular abnormalities were reported in 11/15 individuals; strabismus was most common (6/15) followed by astigmatism (5/15), myopia (3/15), amblyopia (2/15), hyperopia (2/15), and nystagmus (1/15). Sleep disturbance was reported in 10/15 individuals, often characterized by difficulty falling asleep, night wakening, and difficulty falling back asleep. Feeding issues were reported in 10/15 of individuals. The average Body Mass Index (BMI) in our cohort was 17.28 (n=14), which is slightly below the normal BMI range of 18.5 to 24.9; however, children under ten years old (n=10) had an average BMI of 15.3, which is considered severely underweight, whereas children over ten (n=4) had an average BMI of 22.2, which is within the typical BMI range.

Pigmented skin lesions were reported in 8/13 individuals. Recurrent infections were often reported (8/15); the most common was recurrent otitis media (6/15), followed by urinary tract infections (3/15), and respiratory infections (3/15).

Similar dysmorphic features were identified across the cohort (n=14) (Table 3). Notably, all but one individual had malar hypoplasia and a flat midface. A majority of participants had a high arched palate (57%) and a bulbous nose (57%), while 50% had a pointed chin. Other dysmorphic features included nail hypoplasia (46%), wide nasal bridge (43%), long philtrum (43%), full cheeks (29%), and widely spaced teeth (29%).

#### Common findings

Neonatal Intensive Care Unit (NIC)U stays were required for 6/15 individuals (durations ranged from two hours to 15 days, none due to premature birth). Additional neonatal issues gathered through the PDDBI included difficulty latching/feeding (11/13), reflux/spit up (4/13), low birth weight (3/13), needed oxygen at birth (3/13), and irritable/fussy (1/13). Additional pregnancy complications (8/15) included intrauterine growth restriction (3/15), small for gestational age (3/15), gestational diabetes (2/15), nuchal cord thickening (2/15), and oligohydramnios (2/15). The average birth weight was 6lbs 1oz (range: 4lbs 8oz to 7lbs 12oz) and the average gestational period was 38+1 weeks (range: 36 to 40 weeks). The neurological assessment revealed hypertonia in 5/15 individuals, and four of these individuals also had hypotonia. Hearing abnormalities were reported in three individuals. Precocious puberty was present in 2/10 individuals who were over the age of 5.

#### Other less frequent findings

Reported findings include seizures (2/15): one individual had experienced one known seizure episode and was not currently treated with anti-epileptic drugs and a second individual had a history of staring spells, an abnormal electroencephalogram, and ongoing treatment with oxcarbazepine. One individual had a congenital heart defect. Additionally, one participant had both hypothyroidism and obstructive sleep apnea.

### Comparisons Across Variant Classes

An examination of the distribution of neuropsychological profiles as a function of variant class showed evidence for an association of greater severity in clinical phenotype with missense variants/in-frame deletions (Figures 2-4). We carried out further, exploratory analyses in a subset of measures, including Vineland-3 scores, DQ, and ASD diagnosis (Figure 6).

**Figure 6.**
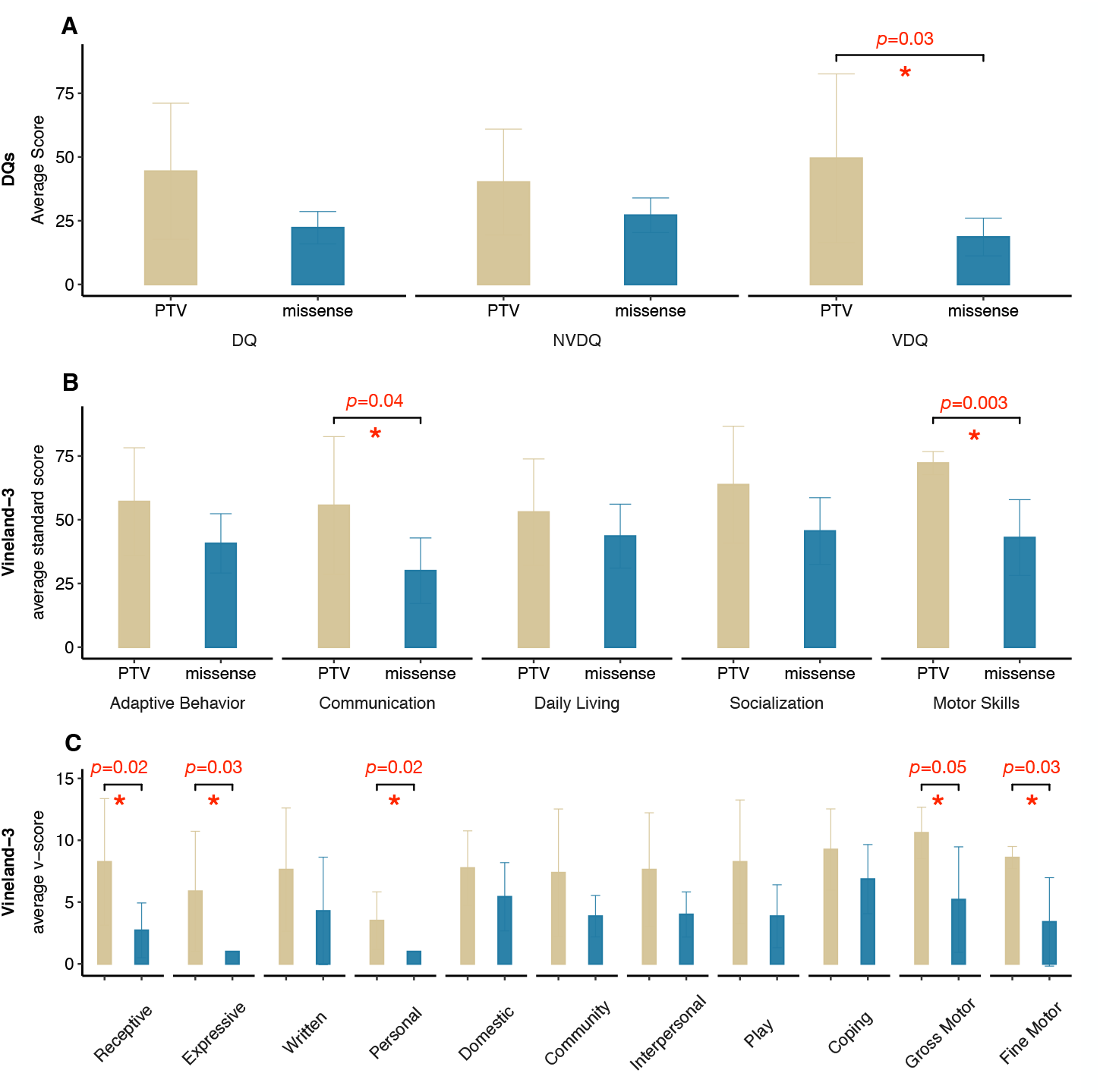
Phenotypic Comparisons Across Variant Classes. A. Average scores for full scale DQ, Nonverbal DQ, and Verbal DQ, comparing the two variant types. B. Average Vineland-3 standard scores, comparing the two variant types. C. Average Vineland-3 v-scale scores, comparing the two variant types. Independent sample t-tests or chi- square analyses were performed to compare the neurobehavioral profiles across variant types. PTV, protein truncating variant; missense, missense variants or in-frame deletions.

The PTV group had significantly higher verbal DQ scores (49.5±33.1, p=.032) when compared to the missense/in-frame deletion group (18.6±7.4) (Figure 6A). With regard to adaptive behavior, assessed via the Vineland-3, individuals in the PTV group showed greater skills in multiple areas, when compared to individuals in the missense variants/in-frame deletions. This was true for the Communication domain (55.6±27.0 versus 30.0±12.9, p=0.037) [and the Expressive Language subdomain (5.9±4.9 versus 1.0±0.0, p=0.025), Receptive Language subdomain (8.38±5.1 versus 2.7±2.2, p=0.020)]. Similarly, the PTV group showed significantly higher scores in the Motor domain (72.2±4.5 versus 43±14.9, p=0.003) [and the Gross Motor subdomain (10.6±2.1 versus 5.2±4.3, p=0.045) and the Fine Motor subdomain (8.6±0.9 versus 3.4±3.6, p=0.029)]. Analyses were also carried out without data from the participant carrying the c.679+3_679+4delinsTT variant, since functional studies have not been done to definitively classify this variant as a PTV. All statistics stayed significant with the exception of the Vineland-3 Communication domain (Table S7). However, both the Receptive and Expressive Language subdomains remained significantly different.

A trend was observed for a lower rate of ASD diagnosis in the PTV group (37.5% versus 85.7%, p=0.057).

On multiple measures, Cohen’s d effect sizes were >0.8 reflecting a large effect (Table S7).

## DISCUSSION

Previous studies of DDX3X syndrome are based largely on chart review of medical histories, with only modest direct real-time assessment of the neurobehavioral and psychiatric manifestations. The literature has mainly focused on ID/DD severity, neurological phenotype (e.g., hypotonia, structural brain abnormalities, epilepsy, movement disorders), and medical comorbidities such as precocious puberty, visual/hearing abnormalities and scoliosis [1, 13, 15, 46] (Table 4). The behavioral phenotype has been noted but not described in detail [1, 15], and ASD, ADHD, and sensory features have not been directly studied. To address this knowledge gap, in the current study we carried out extensive phenotyping, using prospective neuropsychological, neurological, clinical genetic, and psychiatric assessment in 15 individuals with DDX3X syndrome.

**Table 4:** Comparison with past literature

|  | Tang, 2021 | Snijders Blok, 2015 | Wang, 2018 | Beal, 2019 | Lennox, 2020 |
| --- | --- | --- | --- | --- | --- |
| Sample Size (F,M) | 14,1 | 38,0 | 28,0 | 6,0 | 104,3 |
| ID | 80%* | ID or DD<br>100% | ID or DD<br>100% | 50% | ND |
| Motor Delays | 87%* |  |  | 67% | ND |
| Speech Delays | 93%* |  |  |  | ND |
| ASD | 60%* | ASD,<br>hyperactivity,<br>aggression<br>53% | ASD and other<br>behavioral<br>problems<br>21% | ASD,<br>hyperactivity,<br>aggression<br>33% | 21% |
| ADHD | 53%* |  |  |  | 15% |
| Generalized Anxiety Disorder | 7%* | ND | ND | ND | ND |
| Visual Motor Integration abnormalities | 54%* | ND | ND | ND | ND |
| Abnormal expressive language<sup>1</sup> | 80%* | ND | ND | ND | ND |
| Abnormal receptive language<sup>2</sup> | 73%* | ND | ND | ND | ND |
| Sensory symptoms | 100%* | ND | ND | ND | ND |
| Gait Disturbance / Movement Disorders | 100%* | 45% | 61% | 33% | ND |
| Hypotonia | 93%* | 76% | 68% | 0% | 82% |
| Gastrointestinal Abnormalities | 80% | ND | ND | ND | ND |
| Ocular/Vision Abnormalities | 73%* | 34% | 32% | 50% | 36% |
| Sleep Disturbance | 67%* | ND | ND | 33% | ND |
| Abnormal Brain MRI | 86%* | 57% | 90% | 25% | 94%* |
| Corpus Callosum abnormalities | 64%* | 35% | ND | 25% | 87%* |
| PMG / cortical dysplasia | 7%* | 11% | ND | 0% | 12%* |
| Epilepsy/Seizures | 13% | 16% | ND | 17% | 20% |
| Cardiac Defects / Disease | 7% | ND | 71%<sup>3</sup> | 17% | 15% |
| Precocious puberty | 20% | 13% | ND | 0% | 8% |

\*Prospectively assessed
ND, Not Determined
<sup>1</sup> >1.5 SD below the mean on the Expressive Vocabulary Test, Second Edition or could not complete
<sup>2</sup> >1.5 SD below the mean on the Peabody Picture Vocabular Test, Fourth Edition or could not complete
<sup>3</sup> 5 of 7 individuals assessed

In our cohort, consistent with previous reports, *DDX3X* variants clustered in the helicase ATP-binding domain or the helicase-C domain, with the exception of one variant occurring at the start codon. Our cohort included 4 novel variants (p.Glu285Lys, p.Gln241Hisfs*53, p.Gln308*, and p.Arg292Leu) and 11 that were previously identified. Previous literature has suggested that the Arg326His variant has a more severe clinical outcome [13]. This variant has been reported in the literature four times, and all individuals have a history of PMG [13, 16]. Our cohort has a participant with a variant at the same amino acid, Arg326Cys, who also has a history of PMG, further implicating an association between variants at this location and PMG. Additionally, participants with variants at amino acid 415 have been reported to have a more severe clinical presentation. Four individuals have been described with variants at this location, 2 with a history of PMG and 3 with corpus callosum abnormalities and enlarged ventricles [13, 17]. Our participant does not have a history of PMG, however severe developmental delays and medical comorbidities are present.

Cognitive functioning was directly evaluated, and 80% of our cohort met criteria for a DSM-5 diagnosis of ID. We observed a wide range in cognitive ability, which appeared to be associated with the participants’ variant type. Additionally, participants without ASD showed significantly higher DQ scores than participants with ASD. ADHD did not show an association with DQ scores. Adaptive behavior deficits were present in all participants, indicating that even individuals with higher cognitive ability had challenges applying their skills appropriately in daily life.

Language milestones were significantly delayed, and 5 of 15 individuals were non-verbal at the time of assessment. For those in the cohort who were verbal, both receptive and expressive language skills were significantly below age expectations, with expressive language scores showing greater deficits than receptive language scores. The average age of first words occurred at 2.4 years, more than a year after typically expected. Participants without ASD scored significantly higher on all language assessments compared to individuals with ASD.

In addition, behavioral comorbidities were prospectively studied for the first time. DSM-5 diagnoses of ASD were present in 60% of the cohort, and many of the participants who did not meet full ASD criteria exhibited ASD traits such as repetitive behaviors and sensory symptoms. Over half of individuals presented with clinically significant hyperactivity and attention problems and met criteria for ADHD, combined type. Frustration intolerance was also common.

Results from sensory assessments indicated higher rates of sensory hyporeactivity and sensory seeking, compared to typically developing counterparts. Sensory hyporeactivity has been implicated in other genetic syndromes associated with ASD such as Phelan-McDermid syndrome and ADNP syndrome [47, 48] and is often associated with high pain tolerance.

Common medical comorbidities included gastrointestinal difficulties such as constipation and reflux, and recurrent common infections, such as ear, urinary tract, and respiratory tract infections. The ocular phenotype was extended to include features such as astigmatism, myopia, amblyopia, nystagmus, and hyperopia, in addition to strabismus. We replicated evidence of brain abnormalities, microcephaly, and a range of gait abnormalities for those affected by DDX3X syndrome.

Recent analyses of the DDX3X syndrome phenotype provided initial evidence for the association of distinct classes of genetic variants with severity of phenotypes [13]. When we compared phenotypes of individuals with PTV variants to those with missense variants or in-frame deletions, we confirmed that individuals with missense variants, on average, demonstrated more severe clinical phenotypes, including lower intellectual, language, and adaptive functioning. We completed analysis with and without the participant with polymicrogyria and still see significant differences between missense and PTV groups; hence the presence of polymicrogyria, by itself, does not appear to be a simple way of defining the most severe mutations (Table S7). A significant limitation for these analyses is that, with the current sample size, we did not correct for multiple testing. However, for many findings, the Cohen’s d effect sizes were >0.8, indicating that larger sample sizes will continue to show significant differences.

Prospective studies are both expensive and time-consuming, but can have important advantages compared to medical record review. First, such studies can apply gold standard diagnostic instruments which may not be used in typical clinical settings. In addition, all the assessments can be done by a single integrated team which provides more consistent phenotyping and opportunities for consensus diagnoses. From our analyses, a significant proportion of individuals with DDX3X syndrome have definitive diagnoses of ASD and ADHD. Another advantage of prospective approaches is that subclinical assessments and biomarker determination can be part of the phenotyping. In the current report, we include measures such as SRS and a novel sensory biomarker (SAND), demonstrating, for the first time, definitive sensory changes in DDX3X syndrome. Finally, all items and results can be shared via de-identified databases, subject to consent and appropriate compliance review, without requiring additional consent from participants.

### Limitations

The main limitation of our study is the small sample size. Additionally, without functional studies on each of our variants we were unable to group our variants into three potential types (i.e., PTVs, hypomorphic missense, or dominant negative missense variants) [13]. We instead grouped them into two classes (i.e., PTVs, and missense, which would include both hypomorphic and dominant alleles).

### Conclusions

DDX3X syndrome is emerging as a major cause of NDDs in girls. This study uniquely explored the clinical and neuropsychiatric phenotype of individuals with DDX3X syndrome using prospective analyses. Comprehensive clinician-administered evaluations and standardized diagnostic and neuropsychological assessments were administered to characterize the neuropsychiatric dimensions of DDX3X syndrome, including the prevalence of ASD, and to more clearly define language and adaptive functioning in these individuals. Our findings offer a deeper understanding of the behavioral, medical, and developmental challenges that children and adolescents with DDX3X syndrome experience, while informing clinical guidance and potential interventions. As research focused on delineating the clinical phenotypes and natural history of this syndrome continues, more tailored, comprehensive and developmentally-informed assessment and treatment approaches will emerge. These resources will, in turn, support caregivers and clinicians in providing more informed and specific care, treatments, and clinical guidance for the conditions and challenges that affect those with DDX3X syndrome.

Looking ahead, genotype-phenotype correlations in DDX3X syndrome will benefit from robust and uniform phenotypic approaches. In a prospective research setting, specific variants or cases of interest can be over-sampled, which allows exploration of more rare events and/or emergent events that require, or demand, further analyses. The role of potential dominant negative variants in phenotypic expression will be part of our future studies. In addition, the prospective studies reported here will form the basis for longitudinal studies on DDX3X syndrome.

## Data Availability

The majority of the dataset used during the current study is included in this published article and Supplementary File. The remainder of the dataset is available from the corresponding author on reasonable request and may require ethics review.

## List of abbreviations

ADHD: Attention Deficit/Hyperactivity Disorder
ADI: Autism Diagnostic Interview
ADI-R: Autism Diagnostic Interview-Revised
ADOS-2: Autism Diagnostic Observation Schedule, Second Edition
ASD: Autism Spectrum Disorder
BMI: Body Mass Index
CBCL: Achenbach Child Behavior Checklist
DAS-II: Differential Ability Scales-Second Edition
DCDQ: Developmental Coordination Disorder Questionnaire
DD: Developmental Delay
DQ: Developmental Quotient
DSM-5: Diagnostic and Statistical Manual, Fifth Edition ID Intellectual Disability
IQ: Intellectual Quotient
MCDI: MacArthur-Bates Communicative Development Inventory
MRI: Magnetic Resonance Imaging
Mullen: Scales Mullen Scales of Early Learning
NICU: Neonatal Intensive Care Unit
NDDs: Neurodevelopmental Disorders
PDDBI: Pervasive Developmental Disorder Behavior Inventory
PMG: Polymicrogyria
PTV: Protein Truncating Variant
RBS-R: Repetitive Behavior Scale-Revised
SAND: Sensory Assessment for Neurodevelopmental Disorders
SB-5: Stanford-Binet Intelligence Scales, Fifth Edition
SRS-2: Social Responsiveness Scale, Second Edition
SSP: Short Sensory Profile
TD: Typically Developing
Vineland-3: Vineland Adaptive Behavior Scales, Third Edition
VMI-6: Beery-Buktenica Developmental Test of Visual-Motor Integration, Sixth Edition

## Declarations

### Ethics approval and consent to participate

Parents or legal guardians of all participants in this study provided informed consent prior to study participation. This study was approved by the Mouth Sinai Institutional Review Board, Program for the Protection of Human Subjects (Study ID: 98-436).

### Consent for publication

Additional consent was obtained from all parents and guardians for publication of data. Specific consent for all photographs was also obtained.

### Author’s contributions

Conception and design of study: DEG, JDB, PMS, AKol; Acquisition of data: DH, JZ, IGK, JHF, YF, RL, CL, PMS, DEG; Analysis and/or Interpretation: DEG, TL, JHF, LT, CL, PB, JDB, PMS; Drafting the manuscript: DEG, LT, TL SG, JDB; Revising the manuscript for critically for important intellectual content: LT, TL, JHF, SDR, AKos, AKol, JDB, PMS, DEG; Approval of the version of the manuscript to be published: LT, TL, SG, DH, JZ, IGK, JHF, YF, RL, CL, PB, EF, MSB, SDR, AKos, AKol, JDB, PMS, DEG.

## Acknowledgements

The Seaver Autism Center research team would like to thank all of the families affected by DDX3X syndrome, not only for their time, patience, and generous efforts in participating in our research studies, but also for their dedication and commitment to the larger scientific and research community.

## FIGURE LEGENDS

**Figure 5.** Photographs of participants. Photographs of participants.

## Supplementary Information

Table S1: *DDX3X* variants in the cohort

Table S2: Individual scores on neuropsychological evaluation Table S3: Medical comorbidities

Table S4: Brain MRI findings

Table S5: Neurological examination

Table S6: Summary of dysmorphic features

Table S7: Statistics and Cohen’s D for genotype-phenotype analysis

Figure 1. A. Frequency histograms for the Child Behavior Checklist (CBCL) Internalizing and Externalizing composite scales, Depressive and Anxiety scales, and Attention Deficit/Hyperactivity and Defiant DSM-oriented scales for ADHD and oppositional defiant disorder. T-scores have a mean of 50 and standard deviation of 10. B. Frequency histograms for the Vineland-3 Internalizing and Externalizing scales. V scores have a mean of 15 and standard deviation of 3. In all plots, higher scores indicate greater deficits. In panels A & B, distribution of standard scores in typically developing individuals are shown as black lines, together with associated standard deviations (dashed lines). PTV, protein truncating variant; missense, missense variants or in-frame deletions.

